# MyoPath: A Deep Learning Pipeline for Objective Morphometric Assessment of Skeletal Muscle Biopsies

**DOI:** 10.64898/2026.05.27.26349805

**Authors:** Huahua Zhong, Mingshi Gao, Shang Ma, Wenhan Zhang, Nachuan Chen, Kexin Jiao, Bochen Zhu, Jie Song, Chong Yan, Dongyue Yue, Jianying Xi, Wenhua Zhu, Chongbo Zhao, Sushan Luo

## Abstract

Histopathological evaluation of skeletal muscle biopsies relies on subjective, semi-quantitative assessment with no standardized grading system. We developed a four-tissue deep learning segmentation pipeline using Cellpose-SAM for myofiber instance segmentation, a pixel classifier for fat infiltration, and watershed detection for nuclei. We applied this pipeline to 478 H&E whole-slide images from two independent cohorts: HuashanMuscle (n = 79; China; myotonic dystrophy type 1 [DM1], n = 28; limb-girdle muscular dystrophy type R1 [LGMDR1, calpainopathy], n = 12; type R2 [LGMDR2, dysferlinopathy], n = 22; controls, n = 17) and GTEx (n = 399; United States; three-level myopathy spectrum). Thirty-seven unique morphometric features were extracted per sample. Nuclear centralization index (NCI) and fiber size variability coefficient (fiber CV) discriminated myopathy from controls (p = 1.3 × 10^−5^, rank-biserial r = 0.69; and p = 2.9 × 10^−4^, r = 0.58, respectively). DM1 showed the highest NCI (median 0.121), consistent with its centronuclear pathology, and NCI correlated with CTG repeat count (Spearman rho = 0.46, p = 0.042, n = 20). In the GTEx cohort, both biomarkers exhibited significant dose-response trends across the myopathy spectrum (Jonckheere-Terpstra p < 10^−4^). The MyoPath Score, a logistic regression composite of seven pathology indicators trained on GTEx, achieved AUC = 0.788 (LOO-CV 0.735) and transferred to the independent HuashanMuscle cohort with AUC = 0.873 without retraining. Segmentation achieved Dice coefficients of 0.92 (myofiber), 0.95 (fat), 0.87 (nucleus), and 0.88 (connective tissue), with intraclass correlation coefficients exceeding 0.88. NCI and fiber CV provide objective, reproducible quantitative biomarkers for skeletal muscle pathology severity assessment with potential as standardized grading criteria and clinical trial endpoints.

---

Skeletal muscle biopsy is a standard part of the diagnostic workup for patients with neuromuscular symptoms.^1–3^ Interpreting these biopsies requires integrating histomorphologic findings with clinical presentation, and the diagnostic yield depends on the specific clinical question.^2^ Pathological evaluation of hematoxylin and eosin (H&E)-stained sections relies on qualitative descriptors such as “mild fiber size variation,” “occasional centralized nuclei,” and “moderate fatty replacement” that resist standardization across observers and institutions.^1,3^ While pattern-based approaches to muscle biopsy interpretation have been well described,^2^ no internationally accepted quantitative grading system exists for H&E-based skeletal muscle pathology, in contrast to established grading systems in surgical pathology such as the Gleason system for prostatic adenocarcinoma.^4^

The lack of standardized quantification poses particular problems for clinical trials and genotype-phenotype studies. Trials for muscular dystrophies now use histopathological endpoints, but regulatory agencies require objective, reproducible outcome measures.^5,6^ Quantitative magnetic resonance imaging (MRI) can track muscle disease progression,^7,8^ with Dixon fat fraction correlating with CTG repeat length in myotonic dystrophy type 1 (DM1, rho = 0.485) and with clinical severity scales.^8^ However, MRI cannot capture fiber-level features such as nuclear positioning abnormalities, fiber splitting, or fiber-type grouping that carry independent diagnostic significance.

In oncology, computational pathology already automates Ki-67, PD-L1, and mitotic count scoring,^9^ but neuromuscular pathology still depends on manual evaluation. Several tools for skeletal muscle morphometry exist (SMASH,^10^ MyoVision,^11^ MuscleJ,^12^ Open-CSAM^13^), though most address only fiber segmentation and cross-sectional area. Deep learning has more recently been applied to muscle fiber classification^14^ and whole-section quantification with synthetic training data,^15^ but not to multi-layer morphometric analysis of human disease cohorts. Two morphological features lend themselves to automated quantification: nuclear centralization and fiber size variability. In normal muscle, fewer than 3% of fibers have centralized nuclei; above 20%, centronuclear myopathy is pathognomonic.^16^ Fiber size variation, measured as the coefficient of variation of cross-sectional areas, increases in dystrophic and neurogenic processes.^3,17^

In this study, we developed MyoPath, an automated four-tissue segmentation pipeline using Cellpose-SAM^18,19^ within QuPath,^20^ and applied it to 478 H&E whole-slide images from two independent cohorts: HuashanMuscle (n = 79; hereditary myopathies and controls) and GTEx (n = 399; population-based myopathy spectrum).^21^ From 37 extracted morphometric features, we systematically identified seven clinically interpretable pathology indicators and showed that two of them, nuclear centralization index (NCI) and fiber size variability coefficient (fiber CV), serve as primary quantitative biomarkers for muscle pathology severity. We assessed their ability to discriminate disease subtypes, correlate with genotype severity, and transfer across independent cohorts via a composite MyoPath Score.

## Materials and Methods

### Study Design and Ethical Approval

This retrospective, cross-sectional morphometric study analyzed H&E-stained skeletal muscle sections from two independent cohorts. The HuashanMuscle cohort was collected at Huashan Hospital, Fudan University, under institutional review board approval [IRB number 2022-913]. The GTEx cohort was accessed through the Genotype-Tissue Expression Project (dbGaP accession phs000424). All procedures conformed to the Declaration of Helsinki.

### HuashanMuscle Clinical Cohort

The cohort comprised 79 skeletal muscle biopsies from patients with genetically confirmed myopathies and controls with normal histopathology enrolled at Huashan Hospital, Fudan University, Shanghai, China (Table 1). Disease groups included DM1 (n = 28; 15 female; mean age 42.4 ± 10.7 years)^21^, LGMDR1 (calpainopathy, *CAPN3* mutations; n = 12; 4 female; mean age 38.6 ± 13.3 years)^22^, LGMDR2 (dysferlinopathy, *DYSF* mutations; n = 22; 8 female; mean age 32.0 ± 11.6 years)^23^, and controls (n = 17; 10 female; mean age 41.2 ± 15.0 years). Controls were patients who underwent diagnostic muscle biopsy for suspected neuromuscular disease but whose biopsies showed no histopathological abnormalities; they represent diagnostically normal biopsies rather than biologically healthy muscle. The myopathy groups represent moderate-to-severe hereditary muscle disease with established molecular diagnoses. LGMD nomenclature follows the reformed ENMC classification.^24^ Available clinical data included CTG trinucleotide repeat count (DM1, n = 20), grip strength (DM1, n = 12), 10-meter walk test time (DM1, n = 12), and mutation type classification (missense versus loss-of-function) for LGMD patients.

**Table 1.**
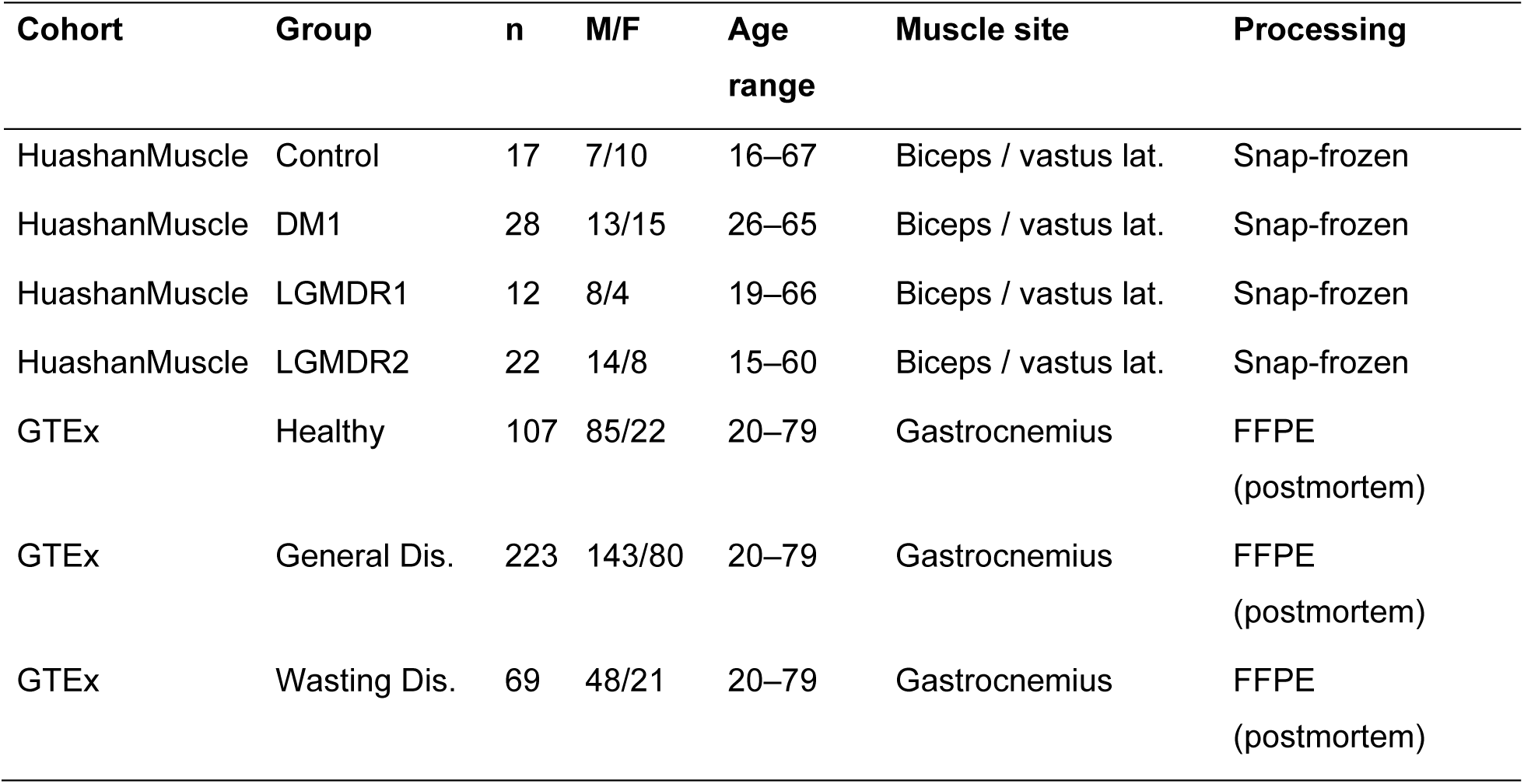
Study cohort characteristics. HuashanMuscle controls were patients with suspected neuromuscular disease whose biopsies showed no histopathological abnormalities. Myopathy groups had established molecular diagnoses (*DMPK* CTG expansion for DM1; biallelic *CAPN3* mutations for LGMDR1; biallelic *DYSF* mutations for LGMDR2). GTEx skeletal muscle samples were collected at autopsy; most skeletal muscle samples in GTEx were harvested from the gastrocnemius region. From 1,001 GTEx donors, 399 with transverse-oriented cross-sections suitable for morphometric analysis were included.

**Table 2.** Segmentation accuracy across four tissue layers. Values are mean +/- SD (n = 30 annotated ROIs from 10 HuashanMuscle samples). Dice and IoU computed by pixel-level comparison between automated segmentation and consensus annotations from two pathologists.

| Tissue layer | Method | Dice coefficient | IoU |
| --- | --- | --- | --- |
| Myofiber | Cellpose-SAM | 0.92 ± 0.03 | 0.85 ± 0.06 |
| Fat | Pixel classifier | 0.95 ± 0.02 | 0.91 ± 0.03 |
| Nucleus | Watershed | 0.87 ± 0.04 | 0.78 ± 0.06 |
| Connective | Boolean subtract | 0.88 ± 0.04 | 0.78 ± 0.06 |

### GTEx Population Cohort

The GTEx project provided H&E-stained whole-slide images of skeletal muscle from 1,001 postmortem donors collected across multiple biobanks in the United States.^25^ Of these, 399 contained transverse-oriented muscle cross-sections suitable for morphometric analysis; the remaining 602 were excluded because of longitudinal sectioning, tangential cutting angle, or insufficient tissue quality. The 399 qualifying samples were classified into a three-level health spectrum based on cause of death as described by our previous work.^26^: healthy (n = 107; accident or suicide, no systemic disease), general diseases (n = 223; various comorbidities at death), and wasting diseases (n = 69; cancer, cachexia, or tuberculosis). The cohort included 276 males and 123 females, aged 20 to 79 years.

### Whole-Slide Image Acquisition

For the HuashanMuscle cohort, open biopsies were obtained from biceps brachii or vastus lateralis muscle. Specimens were snap-frozen in isopentane cooled by liquid nitrogen following standard procedures.^3^ Serial cryosections were cut at 8 µm thickness using a Leica CM 1850 cryostat. Sections were stained with hematoxylin and eosin (H&E) according to routine protocols and digitized using an Olympus VS200 whole-slide scanner at ×40 magnification.

For the GTEx cohort, skeletal muscle samples were collected at autopsy with variable postmortem ischemia intervals documented via the Hardy Scale.^25^ Tissues were formalin-fixed and paraffin-embedded (FFPE) following the GTEx Biospecimen Protocol, and H&E-stained sections were prepared at participating biobanks. Compared with snap-frozen HuashanMuscle biopsies, the FFPE autopsy specimens showed inferior histological preservation, including nuclear morphology distortion and tissue fragmentation, a recognized limitation of postmortem tissue processing that may attenuate morphometric measurements.

### Automated Four-Tissue Segmentation Pipeline

A four-tissue segmentation pipeline was implemented in QuPath^20^ with Cellpose-SAM integration^18,19^ (Fig. 1). To balance segmentation accuracy with computational efficiency, deep learning was reserved for myofiber instance segmentation, the most morphologically challenging task, while conventional image analysis methods were used for the remaining tissue components. This design enables processing of a 1,500 × 1,500 µm ROI in approximately 5 minutes on a single GPU, making whole-slide analysis practical for clinical-scale cohorts.

**Figure 1.**
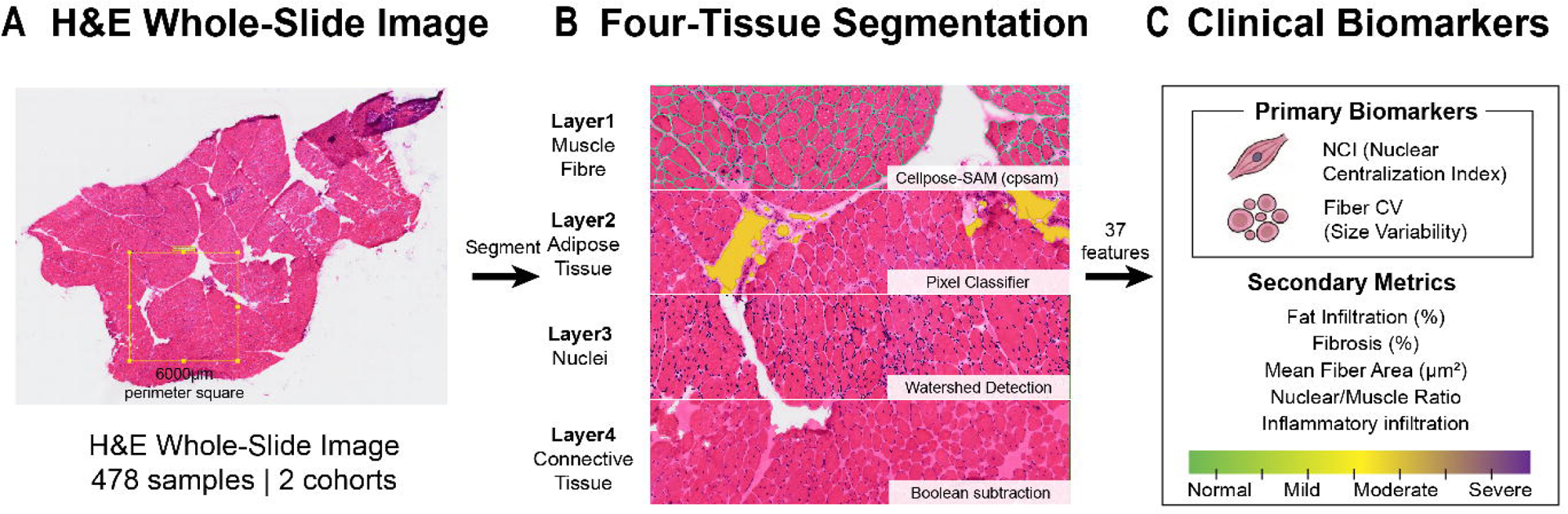
MyoPath pipeline overview. (A) H&E-stained whole-slide image of skeletal muscle with a square region of interest (6,000-µm perimeter). 478 samples from two cohorts were analyzed. (B) Four-tissue segmentation. Layer 1: myofiber instance segmentation using Cellpose-SAM (cpsam); Layer 2: adipose tissue identification using a pretrained pixel classifier; Layer 3: nuclear detection using watershed segmentation on the hematoxylin channel after color deconvolution; Layer 4: connective tissue defined by Boolean subtraction. Deep learning is applied only to myofiber segmentation; the remaining layers use conventional methods for computational efficiency. (C) From 37 extracted morphometric features, two primary biomarkers (nuclear centralization index [NCI] and fiber size variability coefficient [fiber CV]) and five secondary pathology indicators are derived, yielding a severity spectrum from normal to severe.

### Layer 1 (myofiber instances)

Individual myofibers were segmented using the Cellpose-SAM model (cpsam),^19^ a hybrid architecture combining Cellpose’s gradient flow representation with the Segment Anything Model’s pretrained vision transformer backbone.^27^ Parameters were optimized for skeletal muscle cross-sections: expected fiber diameter 100 µm, 4× downsampling, 500-pixel tile overlap, GPU acceleration enabled.

### Layer 2 (fat infiltration)

Adipose tissue was identified using a pretrained QuPath pixel classifier distinguishing fat from myofibers and endomysial connective tissue based on H&E staining intensity. Minimum area threshold was 50 µm^2^.

### Layer 3 (nuclei)

Individual nuclei were detected using QuPath WatershedCellDetection on the hematoxylin channel obtained by color deconvolution of the H&E image (size range 10–500 µm^2^, intensity threshold 0.4, Gaussian sigma 1.5 µm).

### Layer 4 (connective tissue)

Endomysial and perimysial connective tissue was defined as the remaining ROI area after subtracting myofiber, fat, and nuclear regions (connective area = ROI area − muscle area − fat area − nucleus area). This residual approach avoids the need for a separate connective tissue classifier and captures both fibrotic endomysium and perimysial septa. The connective tissue fraction serves as a proxy for fibrosis percentage.

Layers 1–3 produced independent parallel annotations within the same section; Layer 4 was computed from their union. Each tissue component could be analyzed separately or in combination.

### Morphometric Feature Extraction

Thirty-seven unique morphometric features were extracted per sample, comprising raw measurements (tissue areas, fiber size statistics, nuclear counts and positions) and derived pathology indicators. Full definitions with formulas are provided in Supplementary File 1.

From these 37 features, seven were designated as **pathology indicators**, clinically interpretable metrics that correspond to the cardinal features assessed during muscle biopsy evaluation:

1. **Nuclear centralization index (NCI):** the mean relative radial position of nuclei within muscle fibers (range 0–1; 0 = subsarcolemmal, 1 = central). Normal muscle shows NCI < 0.03; elevated values indicate centronuclear myopathy, regeneration, or DM1-associated nuclear mispositioning.^3,16,28^
2. **Fiber size variability coefficient (fiber CV):** the coefficient of variation (standard deviation / mean) of myofiber cross-sectional areas. Normal CV < 0.25; elevated values reflect pathological fiber size heterogeneity from concurrent atrophy and hypertrophy.^3,10,29^
3. **Fiber shape regularity:** mean circularity (4πA/P²; 1.0 = perfect circle). Values < 0.6 indicate irregular fiber morphology from splitting, angular atrophy, or chronic remodeling.^30^
4. **Fat infiltration (%):** fatty tissue as a percentage of ROI area. A late-stage marker of dystrophic or denervation injury.^31,32^
5. **Fibrosis (%):** connective tissue as a percentage of ROI area, reflecting endomysial and perimysial fibrotic remodeling.^33^
6. **Nuclear/muscle ratio:** ratio of nuclei within muscle fibers to total fiber count. Elevated values reflect nuclear proliferation, regeneration, or inflammatory infiltration.^34^
7. **Inflammatory infiltration:** nuclear density in connective tissue (nuclei/mm²). High values suggest inflammatory cell infiltration or active fibrosis.^35,36^

These seven indicators map to five pathological axes: nuclear positioning abnormality (NCI), fiber size dysregulation (fiber CV), fiber morphology distortion (shape regularity), tissue replacement (fat infiltration, fibrosis), and cellular reaction (nuclear/muscle ratio, inflammatory infiltration). Of these, NCI and fiber CV were designated as primary biomarkers because they captured the two axes with the strongest and most consistent discriminative power across both cohorts while representing biologically independent disease mechanisms. NCI was preferred over the simpler central_ratio because it captures the continuous spatial distribution of nuclei rather than a binary threshold, and fiber CV was preferred over raw size statistics because it is dimensionless and comparable across samples with different fiber calibers.

### Segmentation Quality Assessment

Segmentation accuracy was assessed against manual reference annotations generated by two experienced pathologists (M.G. and S.M.) who independently annotated myofibers, fat regions, and nuclei on 30 randomly selected ROIs from 10 HuashanMuscle samples (5 myopathy, 5 control). Inter-observer disagreements were resolved by consensus. Pixel-level Dice coefficients and intersection-over-union (IoU) were computed per tissue layer by comparing automated segmentation to the consensus annotations. Observer agreement between the automated pipeline and each pathologist was quantified by intraclass correlation coefficients (ICC, two-way random, absolute agreement) for the seven pathology indicators. Intra-sample reproducibility was assessed as the coefficient of variation across five non-overlapping regions of interest per sample.

### Statistical Analysis

Non-parametric tests were used throughout. Between-group comparisons used the Mann-Whitney U test or Kruskal-Wallis test with Bonferroni-corrected pairwise comparisons. Effect sizes were reported as rank-biserial correlation (r). Correlations used Spearman rank correlation. Monotonic trends across the GTEx spectrum were assessed by the Jonckheere-Terpstra test for ordered alternatives.

Discriminative power was evaluated by receiver operating characteristic (ROC) analysis. A composite **MyoPath Score** was derived by logistic regression combining all seven pathology indicators, trained on the GTEx cohort (Healthy vs Wasting diseases, n = 176) and independently validated on the HuashanMuscle cohort (Control vs Myopathies, n = 79). The MyoPath Score is defined as:

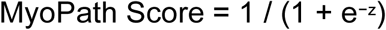

where the coefficients were estimated by L2-regularized logistic regression on z-score standardized features and converted to the raw measurement scale. Leave-one-out cross-validation was performed on the training set to assess overfitting. The same fixed model (coefficients and standardization parameters derived from GTEx) was then applied without retraining to the HuashanMuscle cohort and to the GTEx Healthy-vs-General Diseases comparison. Cross-cohort threshold transferability was evaluated by applying GTEx-derived Youden-optimized thresholds to HuashanMuscle. Analyses were performed in Python 3.12 (scipy 1.12, scikit-learn 1.4). Significance was set at α = 0.05 (two-sided).

## Results

### Segmentation Performance

The MyoPath pipeline processed all 478 whole-slide images (Fig. 1). In a representative HuashanMuscle sample (HE_M3807), the pipeline delineated 953 myofibers, 52 fat regions, and 1,626 nuclei within a square region of interest with a 6,000-µm perimeter (1,500 × 1,500 µm) (Fig. 2A). Dice coefficients for the four tissue layers were 0.92 ± 0.03 for myofiber segmentation, 0.95 ± 0.02 for fat, 0.87 ± 0.04 for nuclei, and 0.88 ± 0.04 for connective tissue (Fig. 2B). Automated measurements of fat infiltration and fibrosis showed strong agreement with human pathologist assessments (Spearman rho = 0.989 and 0.946, respectively; Fig. 2C). Intraclass correlation coefficients exceeded 0.88 for all seven pathology indicators, and intra-sample coefficients of variation were below 0.07 for all metrics.

**Figure 2.**
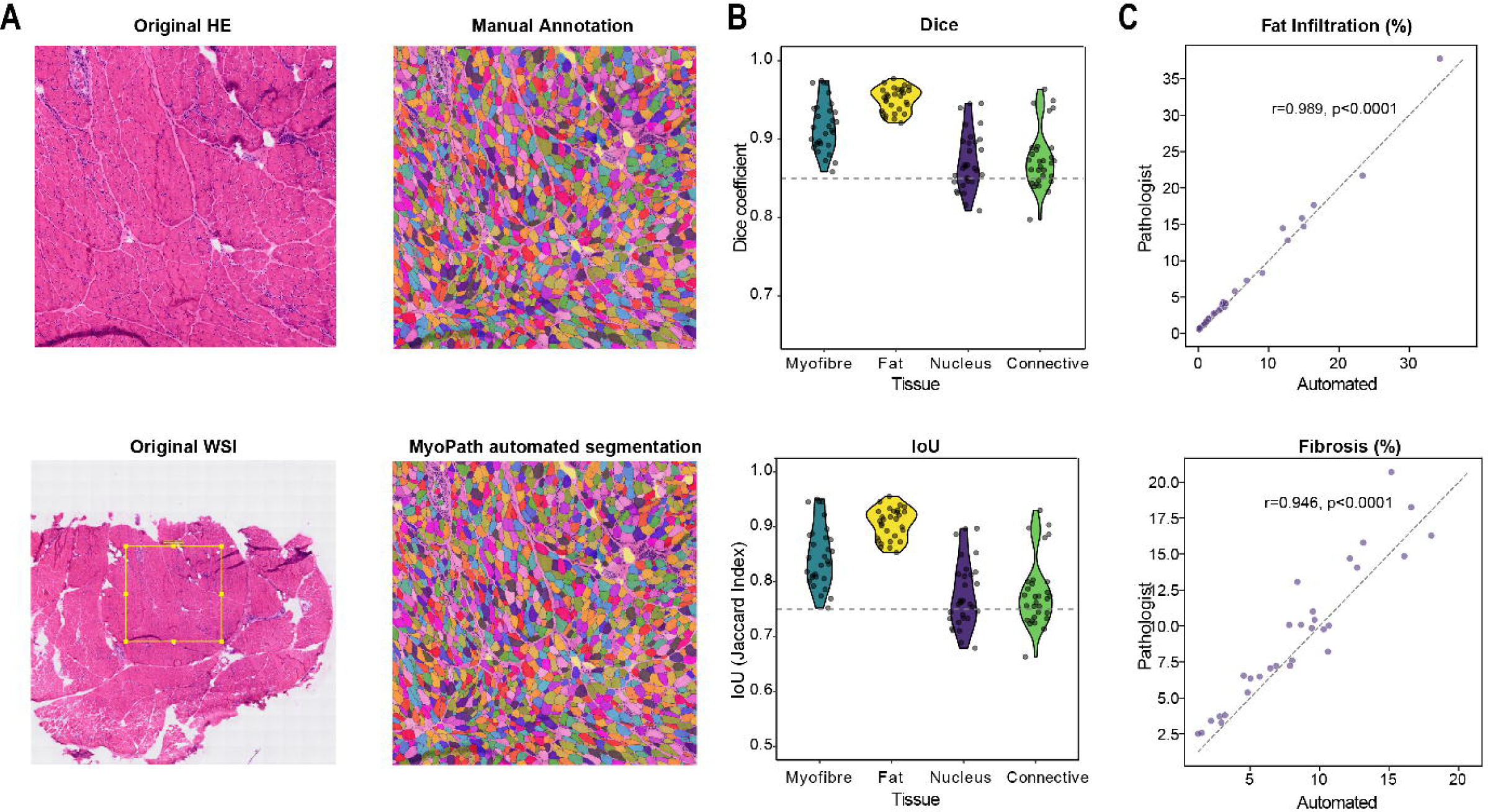
Segmentation validation and observer agreement. (A) Representative sample (HE_M3807). Top row: original H&E image (left) and pathologist manual annotation with each myofiber in a unique color (right). Bottom row: whole-slide image with ROI location (left) and MyoPath automated segmentation (right). (B) Dice coefficients (top) and intersection-over-union (bottom) for four tissue layers (n = 30 annotated ROIs). Dashed lines indicate 0.85 (Dice) and 0.75 (IoU) thresholds. (C) Scatter plots comparing automated measurements with pathologist assessment for fat infiltration (Spearman r = 0.989, p < 0.0001) and fibrosis (r = 0.946, p < 0.0001). Identity lines shown.

### From 37 Features to Seven Pathology Indicators

The pipeline extracted 37 unique morphometric features per sample across tissue composition, fiber size and shape, nuclear distribution, and nuclear localization. All 37 features were screened by univariate Mann-Whitney analysis for discrimination between controls and myopathies (HuashanMuscle) and between healthy and wasting diseases (GTEx).

In the HuashanMuscle cohort (17 controls vs 62 myopathies), 15 of 37 features reached significance (p < 0.05; Fig. 3B). The top-ranked features clustered into three biological categories: nuclear positioning (central ratio AUC = 0.91, NCI AUC = 0.85, peripheral ratio AUC = 0.88), nuclear proliferation (nuclei total count AUC = 0.88, nuclear/muscle ratio AUC = 0.83), and fiber size heterogeneity (fiber CV AUC = 0.79). Tissue composition features (fat infiltration, fibrosis) and fiber shape features did not reach significance (all p > 0.05). In the GTEx cohort (107 healthy vs 69 wasting diseases; Fig. 3C), the ranking shifted: fiber size features rose to the top (fiber area Q1 AUC = 0.75), while nuclear positioning features remained strong (NCI AUC = 0.74, fiber CV AUC = 0.74) and tissue composition features became significant (fat infiltration AUC = 0.67, fibrosis AUC = 0.67). Nuclear proliferation features, which dominated in HuashanMuscle, lost discriminative power in GTEx (nuclear/muscle ratio AUC = 0.50). Features within each category were highly intercorrelated (Spearman |rho| > 0.85 within nuclear positioning; |rho| > 0.80 within fiber size), confirming that a single representative per category captures the information content of multiple redundant metrics.

**Figure 3.**
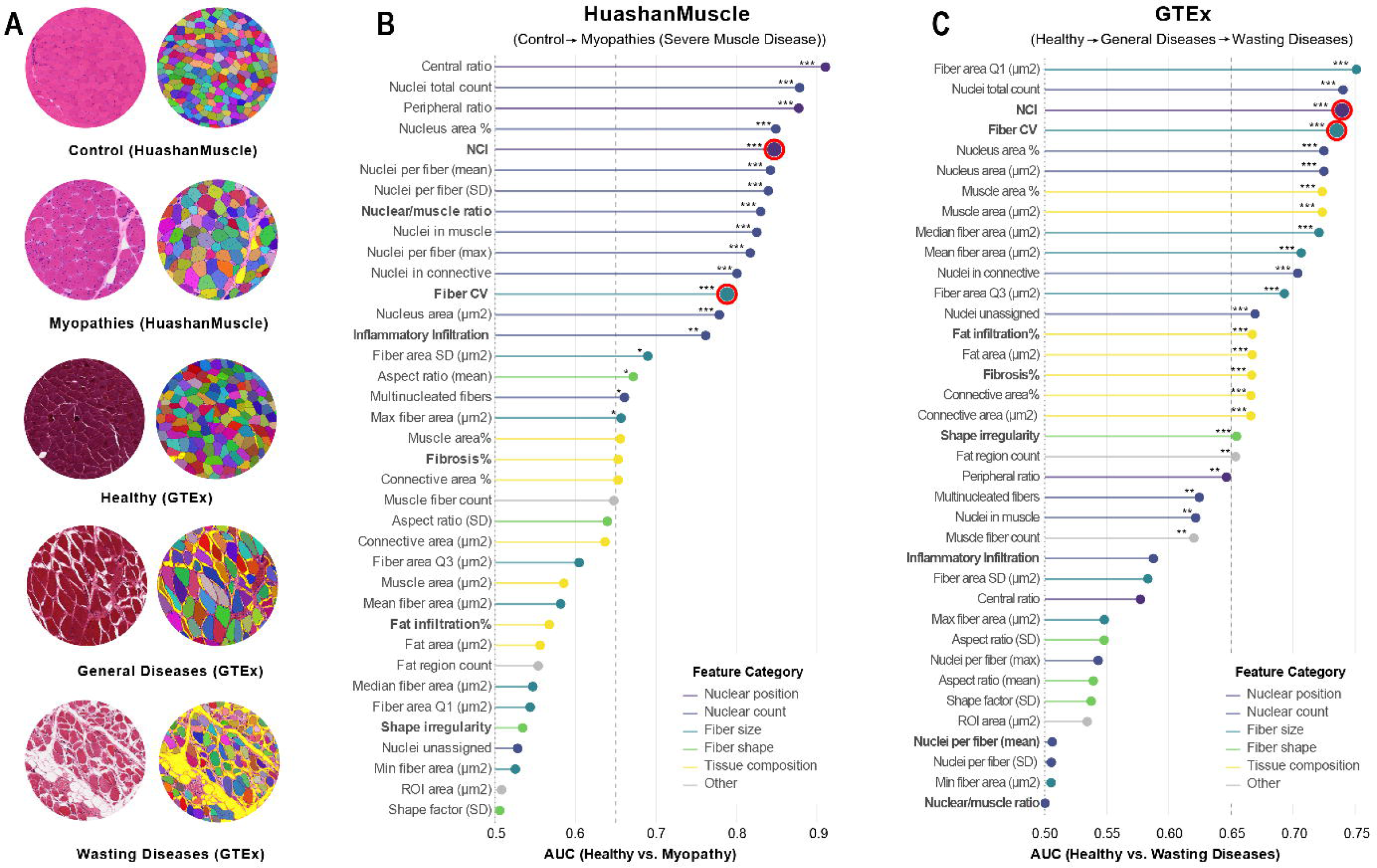
Feature ranking across 37 morphometric features. (A) Representative segmentation results from each diagnostic group. (B) HuashanMuscle cohort: AUC values for discriminating controls from myopathies, ranked by discriminative power. (C) GTEx cohort: AUC values for discriminating healthy donors from those with wasting diseases. Significance: *p < 0.05, **p < 0.01, ***p < 0.001.

Based on these cross-cohort analyses, we selected seven pathology indicators, each representing one clinically relevant dimension: NCI, fiber CV, fiber shape regularity, fat infiltration, fibrosis, nuclear/muscle ratio, and inflammatory infiltration (connective tissue nuclear density). These seven indicators correspond to five pathological axes routinely assessed during muscle biopsy evaluation: nuclear positioning, fiber size dysregulation, fiber morphology distortion, tissue replacement, and cellular reaction.

### Seven Pathology Indicators Discriminate Myopathy from Controls

In the HuashanMuscle cohort, four of the seven pathology indicators significantly differed between controls and myopathies (Fig. 4A). NCI was elevated in myopathy (median 0.077, IQR 0.062–0.121) compared with controls (median 0.049, IQR 0.041–0.058; p = 1.3 × 10^−5^, r = 0.69). Fiber CV was similarly elevated (0.522 vs 0.369; p = 2.9 × 10^−4^, r = 0.58). Nuclear/muscle ratio (4.18 vs 1.80; p = 3.4 × 10^−5^) and inflammatory infiltration (p = 0.003) also differed significantly. Fat infiltration (p = 0.40), fibrosis (p = 0.056), and fiber shape regularity (p = 0.67) did not differ between groups, consistent with the early-to-moderate disease stage of this cohort and the late-stage nature of tissue replacement.

**Figure 4.**
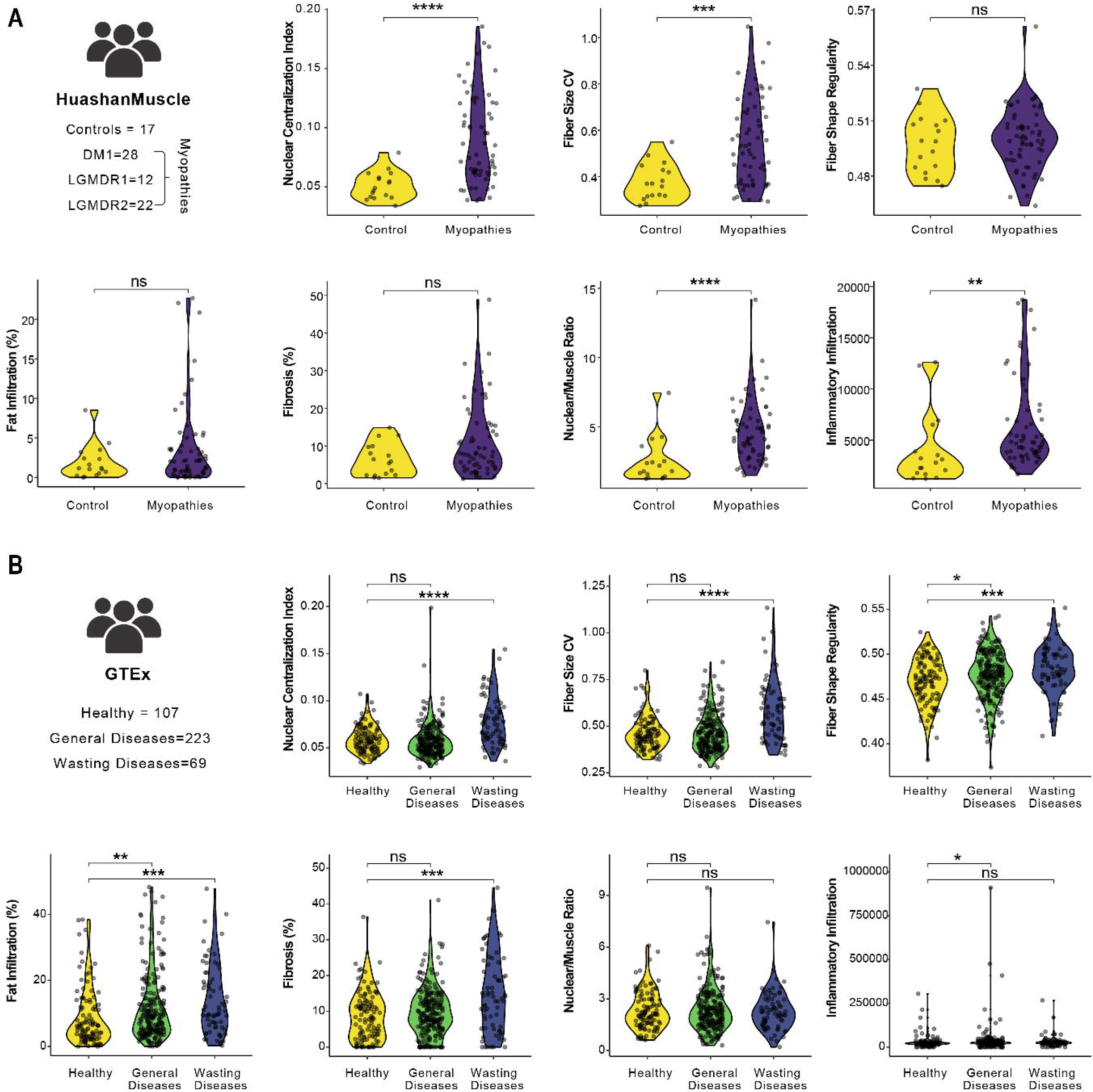
Seven pathology indicators across disease groups. (A) HuashanMuscle cohort: violin plots comparing controls (n = 17) and myopathies (n = 62). (B) GTEx cohort: violin plots across the three-level health spectrum. NCI and fiber CV showed significant monotonic trends (Jonckheere-Terpstra p < 0.0001).

In the GTEx cohort, the seven indicators showed a different pattern across the three-level myopathy spectrum (Fig. 4B). NCI and fiber CV exhibited significant monotonic trends from healthy to wasting diseases (Jonckheere-Terpstra p < 10^−4^ for both). Fat infiltration (p < 0.001) and fibrosis (p < 0.001) also showed significant gradients, reflecting chronic tissue remodeling associated with wasting conditions. Nuclear/muscle ratio did not differ across the spectrum (p > 0.05), consistent with the non-inflammatory nature of disease-related muscle wasting.

Fiber-level indicators dominated in hereditary myopathies, whereas tissue composition indicators contributed more prominently in chronic wasting. This complementary pattern supports the use of all seven indicators as a comprehensive pathological assessment panel, with NCI and fiber CV providing the most consistent discrimination across both disease contexts.

### NCI and Fiber CV as Primary Biomarkers: Clinical Correlations

NCI and fiber CV ranked first and second among the seven pathology indicators in both cohorts. We therefore examined their correlation with clinical measures in DM1 and with mutation severity in LGMD (Fig. 5).

**Figure 5.**
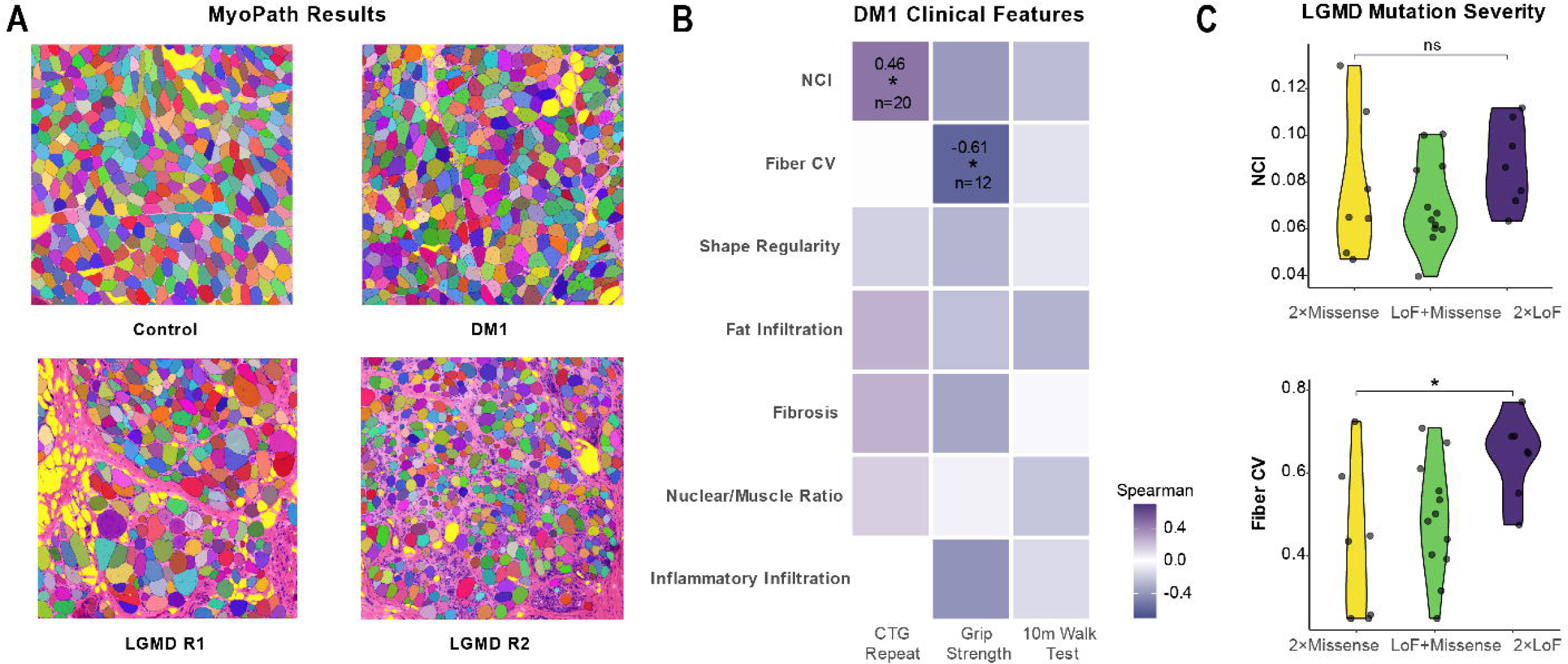
Clinical correlations and mutation severity. (A) Representative MyoPath fiber maps for each diagnostic group. (B) DM1 correlation heatmap. (C) LGMD mutation severity analysis (n = 26).

In DM1 (n = 28), NCI correlated with CTG trinucleotide repeat count (rho = 0.46, p = 0.042, n = 20; Fig. 5B). Expanded CTG repeats in *DMPK* produce toxic RNA foci that disrupt nuclear envelope function,^27^ and the NCI-CTG correlation links automated H&E morphometry to this molecular mechanism. Fiber CV correlated inversely with grip strength (rho = −0.61, p = 0.031, n = 12), indicating that greater fiber size heterogeneity accompanies reduced muscle function. The other five pathology indicators did not correlate significantly with any clinical variable (all p > 0.05).

In LGMD patients with confirmed biallelic mutations (LGMDR1 + LGMDR2, n = 26), fiber CV increased with mutation severity (2× Missense: median 0.44; LoF + Missense: 0.49; 2× LoF: 0.65; Kruskal-Wallis p = 0.044; Fig. 5C). NCI showed a similar trend that did not reach significance (p = 0.23). Fiber size dysregulation may be a more general feature of dystrophic processes than nuclear centralization, which appears more specific to DM1.

### MyoPath Score: Composite Discriminative Power and Cross-Cohort Validation

The MyoPath Score, a logistic regression composite of all seven pathology indicators, was trained on the GTEx cohort (Healthy vs Wasting Diseases, n = 176; Fig. 6). It achieved AUC = 0.788 (LOO-CV = 0.735) in the training set (Fig. 6A), exceeding the best individual indicators (NCI AUC = 0.740; fiber CV AUC = 0.735). NCI and fiber CV carried the largest standardized regression coefficients (β = 0.57 and 0.53), followed by fibrosis (β = 0.39) and shape regularity (β = 0.22).

**Figure 6.**
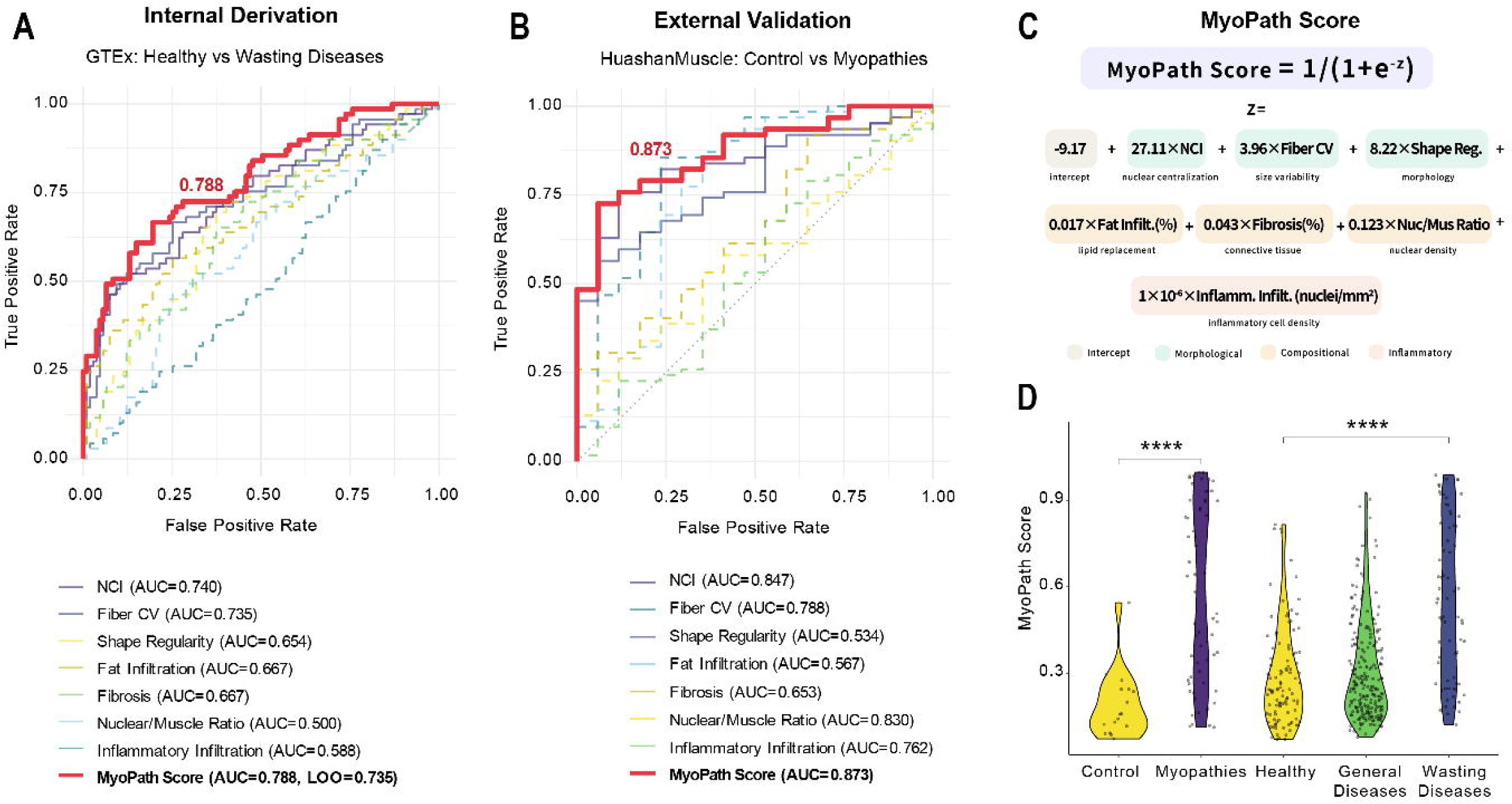
MyoPath Score derivation, cross-cohort validation, and population distribution. (A) Internal derivation: ROC curves for the GTEx training set (AUC = 0.788). (B) External validation on HuashanMuscle (AUC = 0.873). (C) MyoPath Score formula. (D) MyoPath Score distribution across all diagnostic groups (n = 478). Controls and GTEx Healthy donors cluster at low scores (median 0.16 and 0.24), while Myopathies and GTEx Wasting Diseases reach higher scores (median 0.50 and 0.54). Both disease-vs-control comparisons were significant (p < 0.0001, Mann-Whitney).

When applied without retraining to the independent HuashanMuscle cohort, the same model achieved AUC = 0.873 (Fig. 6B). The higher AUC in HuashanMuscle likely reflects the larger phenotypic contrast between genetically confirmed myopathies and histologically normal controls compared with the GTEx wasting-disease gradient. The MyoPath Score formula is shown in Fig. 6C.

Across all 478 samples, the MyoPath Score separated disease groups consistently (Fig. 6D): Controls scored lowest (median 0.16), followed by GTEx Healthy (0.24), GTEx General Diseases (0.25), GTEx Wasting Diseases (0.54), and HuashanMuscle Myopathies (0.50). Both Control-vs-Myopathies and Healthy-vs-Wasting comparisons were significant (p < 0.0001).

## Discussion

Automated morphometric analysis of H&E-stained skeletal muscle can provide objective, reproducible pathology indicators across human skeletal muscle conditions ranging from genetically confirmed hereditary myopathies to subclinical muscle deterioration in a population-based autopsy cohort. NCI and fiber CV separated myopathy from controls, distinguished disease subtypes, and correlated with genetic severity. A composite of all seven indicators, the MyoPath Score, further improved discrimination and generalized across cohorts

Fat infiltration and fibrosis, well-recognized pathological features of chronic myopathy,^2^ proved less discriminative (AUC 0.667 vs 0.740 for NCI). In HuashanMuscle, neither reached significance for discriminating controls from myopathy. Two factors are likely to contribute. First, fat and fibrotic replacement are nonspecific, late-stage changes that also occur with aging, disuse, and obesity.^37^ Second, these tissue-composition metrics are inherently ROI-dependent: their values vary substantially with the location selected for analysis, because fat and connective tissue are often distributed unevenly within a muscle cross-section. A perimysial-rich region will yield higher fibrosis estimates than an endomysial field from the same biopsy. By contrast, NCI and fiber CV are computed per fiber and then averaged, making them less sensitive to ROI placement. This sampling robustness, combined with their sensitivity to early structural changes at the single-fiber level, likely explains why NCI and fiber CV outperformed composition-based metrics across both cohorts. The NCI-CTG correlation (rho = 0.46) links automated H&E morphometry directly to the genetic driver of DM1. Heskamp et al.^8^ showed a similar correlation between MRI fat fraction and CTG repeat length (rho = 0.485); our data add nuclear positioning as a second, independent histomorphometric correlate of repeat expansion severity. That automated H&E fat quantification also agreed with MRI fat fraction (rho = 0.65) supports the biological validity of the measurements.

The GTEx results extend the scope of automated muscle morphometry beyond diagnosed myopathies to general human skeletal muscle health. Fiber atrophy, nuclear centralization, and tissue replacement all tracked a health-status gradient in individuals without clinical neuromuscular diagnosis. The GTEx donors include healthy accident victims, patients with chronic diseases, and those who died from wasting conditions. The histomorphometric changes parallel the transcriptional spectrum we reported previously,^26^ where a continuous Healthy-to-Severe trajectory was identified and validated against clinical features. Our data confirms this spectrum at the tissue level. Population-scale quantitative assessment of muscle pathology, previously impractical by manual methods, is now feasible and may find applications in aging research, sarcopenia, drug-induced myopathy, and clinical trial endpoint assessment.

Several automated muscle morphometry tools have been reported.^10–13^ SMASH^10^ and MyoVision^11^ pioneered open-source fiber analysis; MuscleJ^12^ and Open-CSAM^13^ extended capabilities to centrally nucleated fiber counting and regenerating muscle. More recently, deep learning approaches including FiNuTyper^14^ and the SYNTA synthetic data pipeline^15^ have demonstrated expert-level fiber segmentation. Our pipeline differs in scope: four-tissue segmentation quantifies fiber morphology, fat infiltration, fibrosis, and nuclear distribution from a single H&E section, approximating the information normally obtained from multiple stains. The network-based NDICIA approach of Saez et al.^38^ achieved strong correlation with pathologist severity scoring (R = 0.900) but did not quantify individual fiber-level features. Kabeya et al.^39^ demonstrated CNN-based classification of inflammatory vs hereditary myopathies (AUC 0.996), addressing diagnosis rather than quantitative grading.

The MyoPath Score extends individual biomarker performance by combining all seven pathology indicators into a single composite measure. Trained exclusively on the GTEx population cohort (Healthy vs Wasting Diseases, n = 176), the score achieved AUC = 0.788 with leave-one-out cross-validation AUC = 0.735, indicating modest overfitting and reasonable internal generalizability. More importantly, the same fixed model transferred to the independent HuashanMuscle cohort (AUC = 0.873), which differed from the training set in tissue preparation (snap-frozen vs FFPE), disease etiology (hereditary myopathy vs wasting-related muscle loss), and patient population. This cross-cohort performance gain likely reflects the greater phenotypic contrast between genetically confirmed myopathies and histologically normal controls in HuashanMuscle, compared with the more continuous wasting-disease gradient in GTEx. The standardized regression coefficients confirm that NCI and fiber CV anchor the composite (β = 0.57 and 0.53), with fibrosis contributing a secondary structural component (β = 0.39), consistent with their individual discriminative rankings.

This study has several limitations. The GTEx spectrum classification is based on cause of death rather than clinical muscle diagnosis, introducing misclassification noise that likely attenuates estimated discriminative power; the true effect sizes may be larger than reported. The HuashanMuscle cohort derives from a single center, and sample sizes for certain subgroup analyses were limited (grip strength, n = 12; LGMD mutation subgroups, n = 3–9). GTEx autopsy specimens used FFPE tissue processing with variable postmortem ischemia times, whereas HuashanMuscle used snap-frozen cryosections; the successful cross-cohort transfer of the MyoPath Score despite this difference in tissue preparation suggests robustness, but prospective validation on standardized specimens is warranted. Tissue-composition indicators (fat infiltration, fibrosis) are sensitive to ROI placement within the biopsy, and values from a single ROI may not represent the entire section; future versions should incorporate multi-ROI sampling or whole-section analysis. The pipeline requires QuPath with GPU-enabled Cellpose, though the complete workflow is freely available (https://github.com/Hirriririir/MyoPath).

In summary, MyoPath extracts 37 morphometric features from routine H&E sections, distills them into seven pathology indicators, and identifies NCI and fiber CV as primary biomarkers that discriminate disease subtypes, correlate with genetic severity, and transfer across independent cohorts. The MyoPath Score combines all seven indicators into a single composite measure validated in an external cohort. Multi-center prospective studies are needed, but these biomarkers have potential as standardized grading criteria for skeletal muscle pathology, applicable to hereditary myopathies, aging, sarcopenia, drug-induced toxicity, and disease-related wasting.

## Acknowledgments

The results published here are in part based upon data generated by the Genotype-Tissue Expression (GTEx) Project. We thank the GTEx Consortium for providing open access to whole-slide images of skeletal muscle tissue.

## Author Contributions

H.Z. and M.G. contributed equally to this work. H.Z.: conceptualization, slide preparation, data collection, data analysis, writing—original draft. M.G.: slide preparation, pathological annotation, data collection, writing—original draft. S.M. and W.Z.: data collection, data analysis. N.C., K.J., B.Z., J.S., C.Y., D.Y., J.X., and W.Z.: data collection. C.Z. and S.L.: conceptualization, resources, supervision, funding acquisition. All authors reviewed and edited the manuscript.

## Data Availability

The GTEx data are available through dbGaP (accession phs000424). HuashanMuscle H&E whole-slide images and MyoPath analysis results are available at https://myotoolkit.huashanmuscle.com. The MyoPath analysis pipeline, including the four-tissue segmentation workflow and all analysis scripts, is available at https://github.com/Hirriririir/MyoPath. Additional data is available from the corresponding authors upon reasonable request.

## Declaration of Generative AI and AI-assisted Technologies in the Writing Process

During the preparation of this work the author(s) used the tool ChatGPT (OpenAI) in order to improve readability and language in selected paragraphs. After using this tool/service, the authors reviewed and edited the content as needed and take full responsibility for the content of the publication.

