## Supplementary File 1 for "MyoPath: A Deep Learning Pipeline for Objective Morphometric Assessment of Skeletal Muscle Biopsies"

This supplementary file contains detailed feature definitions referenced in the main text.

---

### 1. Complete Feature List (37 Unique Features)

#### 1.1 Seven Pathology Indicators

These 7 features are clinically interpretable indicators corresponding to cardinal features assessed during muscle biopsy evaluation.

| # | Feature | Unit | Normal range | What it measures |
|---|---|---|---|---|
| 1 | `nuclear_centralization_index` | 0–1 | < 0.03 | Average radial position of nuclei within myofibers. In healthy muscle, nuclei sit beneath the sarcolemma (score near 0). Elevated values indicate nuclei have migrated toward the fiber center, a hallmark of centronuclear myopathies and DM1. |
| 2 | `fiber_cv` | dimensionless | < 0.25 | Coefficient of variation of myofiber cross-sectional areas. Low values indicate uniform fiber size. Elevated values reflect a mixture of atrophic and hypertrophic fibers, characteristic of dystrophic and neurogenic processes. |
| 3 | `fiber_shape_irregularity` | 0–1 | > 0.7 | Mean circularity of fiber cross-sections (1.0 = perfect circle). Low values indicate fibers have lost their normal polygonal shape due to splitting, angular atrophy, or chronic remodeling. |
| 4 | `fat_infiltration_pct` | % | < 5% | Percentage of the ROI occupied by adipose tissue. Reflects fatty replacement of myofibers, a late-stage marker seen in advanced dystrophies and denervation. ROI-dependent. |

| # | Feature | Unit | Normal range | What it measures |
|---|---|---|---|---|
| 5 | `fibrosis_pct` | % | < 10% | Percentage of ROI occupied by connective tissue (computed as ROI minus muscle minus fat). Reflects endomysial and perimysial fibrotic proliferation. ROI-dependent. |
| 6 | `nuclear_muscle_ratio` | dimensionless | 1–3 | Ratio of nuclei located within myofibers to total fiber count. Elevated values reflect nuclear proliferation associated with regeneration, inflammation, or increased satellite cell activity. |
| 7 | `connective_nuclear_density_per_mm2` | nuclei/mm² | < 2,000 | Number of nuclei per square millimeter within connective tissue regions. High values suggest inflammatory cell infiltration or active fibroblast proliferation within the endomysium and perimysium. |

#### 1.2 Underlying Descriptive Features (30 Features)

#### A. Tissue Composition (10 features)

| # | Feature | Unit | What it measures |
|---|---|---|---|
| 8 | `roi_area_um2` | um² | Total area of the analyzed region of interest. Serves as the denominator for percentage-based metrics. |
| 9 | `muscle_fibers_count` | count | Total number of individual myofibers detected by Cellpose-SAM within the ROI. |
| 10 | `muscle_area_um2` | um² | Sum of cross-sectional areas of all detected myofibers. |
| 11 | `muscle_area_pct` | % | Myofiber area as a fraction of ROI area. Decreases with muscle wasting, fat replacement, or fibrosis. |
| 12 | `fat_regions_count` | count | Number of discrete adipose regions identified by the pixel classifier. |
| 13 | `fat_area_um2` | um² | Total area occupied by adipose tissue, excluding overlap with muscle fiber annotations. |
| 14 | `connective_area_um2` | um² | Area of connective tissue, computed by subtracting muscle and fat from total ROI area. |
| 15 | `connective_area_pct` | % | Connective tissue as a fraction of ROI area. Related to but not identical to fibrosis_pct. |
| 16 | `nucleus_area_um2` | um² | Total area occupied by all detected nuclei. |

| # | Feature | Unit | What it measures |
| --- | --- | --- | --- |
| 17 | `nucleus_area_pct` | % | Nuclear area as a fraction of ROI area. Elevated in conditions with increased cellularity. |

#### B. Fiber Size (7 features)

| # | Feature | Unit | What it measures |
| --- | --- | --- | --- |
| 18 | `fiber_mean_area_um2` | um² | Average cross-sectional area of all detected fibers. Decreased in atrophy, increased in hypertrophy. |
| 19 | `fiber_median_um2` | um² | Median fiber area. Less sensitive to outliers than the mean; useful for skewed distributions. |
| 20 | `fiber_std_um2` | um² | Standard deviation of fiber areas. High values indicate heterogeneous fiber sizes. |
| 21 | `fiber_min_um2` | um² | Area of the smallest detected fiber. Very small values may indicate severe atrophy or segmentation artifacts. |
| 22 | `fiber_max_um2` | um² | Area of the largest detected fiber. Very large values may indicate compensatory hypertrophy. |
| 23 | `fiber_q1_um2` | um² | 25th percentile of fiber areas. Marks the lower end of the size distribution; sensitive to grouped atrophy. |
| 24 | `fiber_q3_um2` | um² | 75th percentile of fiber areas. Marks the upper end of the size distribution. |

#### C. Fiber Shape (3 features)

| # | Feature | Unit | What it measures |
| --- | --- | --- | --- |
| 25 | `shape_factor_std` | dimensionless | Standard deviation of circularity across fibers. High values indicate heterogeneous fiber shapes within a sample. |
| 26 | `aspect_ratio_mean` | dimensionless | Mean ratio of bounding box length to width (1.0 = circular, >2.0 = elongated). Detects fiber elongation from tangential sectioning or splitting. |
| 27 | `aspect_ratio_std` | dimensionless | Standard deviation of aspect ratio. High values indicate a mix of round and elongated fibers. |

#### D. Nuclear Distribution (7 features)

| # | Feature | Unit | What it measures |
| --- | --- | --- | --- |
| 28 | `nuclei_total_count` | count | Total number of nuclei detected by watershed segmentation within the ROI (all tissue compartments combined). |
| 29 | `nuclei_in_muscle` | count | Nuclei whose centroids fall within myofiber polygons. Includes subsarcolemmal and internal nuclei. |

| # | Feature | Unit | What it measures |
|---|---|---|---|
| 30 | nuclei_in_connective | count | Nuclei located in connective tissue regions. Includes fibroblasts, inflammatory cells, and satellite cells outside the basal lamina. |
| 31 | nuclei_unassigned | count | Nuclei that could not be assigned to muscle or connective tissue. May include nuclei in fat regions or at tissue boundaries. |
| 32 | nuclei_per_fiber_mean | count/fiber | Average number of nuclei per myofiber. Healthy muscle typically has 1–3 nuclei per cross-sectional profile. |
| 33 | nuclei_per_fiber_std | count/fiber | Standard deviation of nuclei per fiber. High values indicate uneven nuclear distribution across fibers. |
| 34 | nuclei_per_fiber_max | count | Maximum number of nuclei found in any single fiber. Very high values may indicate giant fibers or nuclear clumps. |

#### E. Nuclear Localization (3 features)

| # | Feature | Unit | What it measures |
|---|---|---|---|
| 35 | peripheral_ratio | dimensionless | Fraction of analyzed nuclei in the outer 30% of fiber radius. In healthy muscle this value approaches 1.0, as nuclei are subsarcolemmal. |
| 36 | central_ratio | dimensionless | Fraction of analyzed nuclei in the inner 30% of fiber radius. Values above 0.05 suggest central nuclear pathology. |
| 37 | multinucleated_fiber_count | count | Number of fibers containing more than one nucleus in cross-section. Elevated in regenerating muscle and certain congenital myopathies. |

### 2. Notation and Coordinate System

| Symbol | Definition |
|---|---|
| A_ROI | Total area of the region of interest (ROI) in μm² |
| N_f | Total number of detected muscle fibers |
| a_i | Area of the i-th muscle fiber in μm² |
| p_i | Perimeter of the i-th muscle fiber in μm |
| N_nuc | Total number of detected nuclei |
| s | Pixel size (scanner-dependent, default 0.137 μm/pixel) |
| d | Downsample factor applied during segmentation |

All spatial coordinates originate from QuPath image metadata. Pixel-level measurements are converted to physical units:

$$A\ (\mu m^2) = A\_pixels \times (s \times d)^2$$

$$L\ (\mu m) = L\_pixels \times (s \times d)$$

---

### 3. Tissue Composition Metrics

#### 3.1 Muscle Area

$$A\_muscle = \Sigma\, a_i \quad (i = 1 \text{ to } N\_f)$$

$$muscle\_area\_pct = (A\_muscle \,/\, A\_ROI) \times 100\%$$

#### 3.2 Fat Infiltration

Fat regions are detected by a pre-trained pixel classifier. To avoid double-counting tissue where fat and muscle annotations overlap, the effective fat area is computed as:

$$A\_fat = \Sigma\, area(F_j \setminus \cup M_i) \quad [geometric\ difference]$$

$$fat\_infiltration\_pct = (A\_fat \,/\, A\_ROI) \times 100\%$$

where $F_j$ is the j-th fat polygon, $M_i$ is the i-th muscle fiber polygon, and \ denotes the geometric difference operator.

#### 3.3 Fibrosis (Connective Tissue)

Connective tissue area is computed by Boolean subtraction of muscle and fat from the total ROI:

$$A\_connective = A\_ROI - A\_muscle - A\_fat$$

$$fibrosis\_pct = (A\_connective \,/\, A\_ROI) \times 100\%$$

#### 3.4 Nucleus Area

$$nucleus\_area\_pct = (\Sigma\, a_k^{nuc} \,/\, A\_ROI) \times 100\%$$

---

### 4. Muscle Fiber Size Metrics

#### 4.1 Descriptive Statistics

$$fiber\_mean\_area\_um2 = \bar{a} = (1/N\_f) \times \Sigma\, a_i$$

$$fiber\_std\_um2 = \sigma_a = \sqrt{[\,(1/(N\_f-1)) \times \Sigma(a_i - \bar{a})^2\,]}$$

$$fiber\_median\_um2 = median(a_1, a_2, \ldots, a\_\{N\_f\})$$

$$fiber\_q1\_um2 = Q_1(a), \quad fiber\_q3\_um2 = Q_3(a)$$

$$fiber\_min\_um2 = min(a_i), \quad fiber\_max\_um2 = max(a_i)$$

#### 4.2 Fiber Size Variability (Coefficient of Variation)

$$fiber\_cv = fiber\_size\_variability\_cv = \sigma_a \,/\, \bar{a}$$

| Range | Interpretation |
|---|---|
| < 0.25 | Normal |
| 0.25 – 0.40 | Mild variability |
| 0.40 – 0.60 | Moderate variability |
| > 0.60 | Severe variability |

#### 4.3 Equivalent Diameter

$$d_i^{eq} = \sqrt{(4a_i / \pi)}$$

---

### 5. Muscle Fiber Shape Metrics

#### 5.1 Shape Factor (Circularity)

$$shape\_factor\_i = 4\pi\, a_i / p_i^{\,2}$$

$$shape\_factor\_mean = fiber\_shape\_irregularity = (1/N\_f) \times \Sigma\, shape\_factor\_i$$

A perfect circle yields a shape factor of 1.0. Values below 0.6 indicate irregular fiber morphology; values below 0.4 indicate severely irregular fibers.

#### 5.2 Aspect Ratio (Elongation)

For the bounding box of each fiber polygon with width $w_i$ and height $h_i$:

$$aspect\_ratio\_i = max(w_i, h_i) / min(w_i, h_i)$$

$$aspect\_ratio\_mean = (1/N\_f) \times \Sigma\, aspect\_ratio\_i$$

A value of 1.0 indicates a circular cross-section; values > 2.0 indicate elongated fibers.

---

### 6. Nuclear Distribution Metrics

#### 6.1 Nucleus Assignment

Each detected nucleus centroid $c_k$ is classified by spatial containment testing (Shapely contains):

$$location(k) = muscle \quad if\ \exists\ i: M_i \ni c_k$$

$$connective \quad if\ c_k \in (A\_ROI \setminus \cup M_i \setminus \cup F_j)$$

$$unassigned \quad otherwise$$

$$N\_nuc\text{^}muscle = \#\{k : location(k) = muscle\}$$

$$N\_nuc\text{^}conn = \#\{k : location(k) = connective\}$$

#### 6.2 Nuclei per Fiber

For each fiber $M_i$, the number of contained nuclei:

$$n_i = \#\{k : M_i \ni c_k\}$$

$$nuclei\_per\_fiber\_mean = (1/N\_f) \times \Sigma\, n_i$$

$$multinucleated\_fiber\_count = \#\{i : n_i > 1\}$$

#### 6.3 Nuclear/Muscle Ratio

$$nuclear\_muscle\_ratio = N\_nuc\text{^}muscle / N\_f$$

#### 6.4 Connective Tissue Nuclear Density

$$connective\_nuclear\_density\_per\_mm^2 = N\_nuc\text{^}conn / (A\_connective / 10^6)$$

where A_connective is in $\mu m^2$ and $10^6$ converts to $mm^2$.

---

### 7. Nuclear Centralization Index (NCI)

NCI quantifies the radial position of nuclei within muscle fibers, serving as the primary biomarker for central nuclear myopathy.

#### 7.1 Relative Radial Position

For each nucleus k within fiber $M_i$:

$$d_k^{boundary} = min\_\{p \in \partial M_i\} \|c_k - p\|$$

$$r_i^{eq} = \sqrt{(a_i / \pi)}$$

$$\rho_k = min( d_k^{boundary} / r_i^{eq},\ 1.0 )$$

where $d_k^{boundary}$ is the Euclidean distance from the nucleus centroid to the nearest point on the fiber boundary, $r_i^{eq}$ is the equivalent circle radius, and $\rho_k \in [0, 1]$ ($\rho_k = 0$ at boundary, $\rho_k = 1$ at center).

#### 7.2 Aggregate NCI

$$NCI = nuclear\_centralization\_index = (1/N\_analyzed) \times \Sigma \rho_k$$

where N_analyzed is the number of nuclei successfully localized within muscle fibers.

#### 7.3 Peripheral and Central Ratios

Nuclei are classified into three zones based on their relative radial position:

$$zone(k) = peripheral \quad if\ \rho_k \leq 0.3$$

$$intermediate \quad if\ 0.3 < \rho_k < 0.7$$

$$central \quad if\ \rho_k \geq 0.7$$

$$peripheral\_ratio = \#\{k : \rho_k \leq 0.3\} / N\_analyzed$$

$$central\_ratio \quad = \#\{k : \rho_k \geq 0.7\} / N\_analyzed$$

| NCI Range | Interpretation |
|---|---|
| < 0.03 | Normal (subsarcolemmal nuclei) |
| 0.03 – 0.10 | Mild centralization |
| 0.10 – 0.20 | Moderate centralization |
| > 0.20 | Severe centralization |
